# A metabolome-wide Mendelian randomization study identifies dysregulated arachidonic acid synthesis as a potential causal risk factor for bipolar disorder

**DOI:** 10.1101/2024.02.18.24302998

**Authors:** David Stacey, Beben Benyamin, S Hong Lee, Elina Hyppönen

## Abstract

**Background:** Bipolar disorder (BPD) is a debilitating mood disorder with an unclear aetiology. A better understanding of the underlying pathophysiological mechanisms will help to identify novel targets for improved treatment options and prevention strategies. In this metabolome-wide Mendelian randomization study, we screened for metabolites that may have a causal role in BPD.

**Methods:** We tested a total of 913 circulating metabolite exposures assessed in 14,296 Europeans using a mass spectrometry-based platform. For the BPD outcome, we used summary data from the largest and most recent genome-wide association study (GWAS) to date, including 41,917 BPD cases.

**Results:** We identified 33 metabolites associated with BPD (*p*_*adj*_ <5.48×10^−5^). Most of them were lipids, including arachidonic acid (*β*=−0.154, se=0.023, *p*=3.30×10^−11^), a polyunsaturated omega-6 fatty acid, along with several complex lipids containing either an arachidonic or a linoleic fatty acid side chain. These associations did not extend to other closely related psychiatric disorders like schizophrenia or depression, though they may be involved in the regulation of lithium response. These lipid associations were driven by genetic variants within the *FADS1/2/3* gene cluster, which is a robust BPD risk locus encoding a family of fatty acid desaturase enzymes responsible for catalysing the conversion of linoleic into arachidonic acid. Statistical colocalization analyses indicated that 27 of the 33 metabolites share the same genetic aetiology with BPD at the *FADS1/2/3* cluster, demonstrating that our findings are not confounded by linkage disequilibrium.

**Conclusions:** Overall, our findings support the notion that ARA and other polyunsaturated fatty acids may represent potential targets for BPD.

## Introduction

Bipolar disorder (BPD) is a debilitating mood disorder characterized by recurring episodes of mania and depression with a lifetime prevalence of ~2% (1). Although the clinical features of BPD can vary widely, there are two broad diagnostic subtypes: BPD type I and type II. While episodes of mania are the defining feature of BPD type I patients, BPD type II patients experience hypomanic episodes, which are generally shorter-lived and less debilitating than manic episodes. Furthermore, diagnosis of BPD type II requires at least one major depressive episode, which is not the case for BPD type I. The gold standard drug treatment for BPD is lithium, which has demonstrated unique effectiveness in managing both manic and depressive episodes (2). However, there is considerable variability in the magnitude of response to lithium across BPD patients, with ~30% experiencing complete remission and the remainder responding either partially or not at all (3).

While the exact causes of BPD remain unclear, twin and family studies have shown that BPD is highly heritable, with estimates ranging between 70-90% (4). Accordingly, recent genome-wide association studies (GWAS) have now identified several dozen independent BPD risk loci at genome-wide significance (*p*<5×10^−8^) (5). However, the precise causal genes and mechanisms underlying most of these BPD-risk loci are not yet known.

Molecular quantitative trait locus (molQTL) data have proven instrumental in identifying genes and other molecules regulated by disease-associated loci, though it is often difficult to establish whether they play a causal role in the disease. To address this, Mendelian randomization (MR) – a method that utilizes genetic instruments as proxies for exposures of interest to derive evidence for underlying causality – has been used to highlight several molecular traits as potential causal risk factors for psychiatric disorders (e.g., C-reactive protein and schizophrenia) (6). Moreover, highly scalable MR methods, like generalized summary-data-based MR (GSMR) (7), have emerged that enable MR screens of transcriptomic, proteomic, and metabolomic data.

Since people with psychiatric disorders have, on average, a higher risk of cardiometabolic dysfunction relative to the general population (8, 9), several metabolome-wide MR (MWMR) studies of psychiatric outcomes have been conducted recently. A two-sample MR study using summary data for 486 serum metabolite exposures and five major psychiatric disorders – including BPD – highlighted glycerolipid metabolism as potentially relevant for BPD (10). A similar study of 92 metabolite exposures and eight psychiatric disorders identified glycine, 1-arachidonoylglycerophosphocholine, and glycoproteins as putative BPD protective factors (11). More recently, a study in UK biobank (UKBB) participants with data for 249 circulating metabolites highlighted potential causal roles for polyunsaturated fatty acids (PUFAs), particularly omega-3 PUFAs, in major depressive disorder (MDD) (12).

These findings support the notion that circulating metabolites reflect an important aetiological role in BPD and other psychiatric disorders. Here, to extend on these findings, we have performed a two-sample MWMR analysis of BPD using the most up-to-date publicly available summary data. By incorporating >900 plasma metabolites as exposures (13), our study provides almost twice the metabolomic coverage compared to previous MWMR studies conducted in the psychiatric field (10, 11).

## Methods and Materials

### GWAS summary statistics

For this study we utilized publicly available summary data from several sources. To derive metabolomic exposures, we utilized summary statistics from a multi-trait GWAS of 913 plasma metabolites (**Table S1**) quantified in 14,296 European individuals using the Metabolon HD4 platform (13). For the BPD outcome, we downloaded summary data from the Psychiatric Genetics Consortium (PGC) website, which represents the most recent and largest BPD GWAS available at the time of writing. This dataset comprised up to 413,466 Europeans, including 41,917 BPD cases (5). We also utilized other European GWAS summary datasets for replication and follow-up analyses, including an earlier multi-trait GWAS of circulating metabolites (14) as well as GWAS of several additional psychiatric traits (see **Supplementary Methods**).

### Generalized Summary-data-based Mendelian Randomization (GSMR)

We applied GSMR(7) as implemented in the Genome-wide Complex Trait Analysis (GCTA) suite of tools (v1.94.1) (15). We filtered out variants with discordant allele frequencies across datasets (--diff-freq 0.2). To select variants for LD clumping as part of the GSMR workflow, we specified a significance threshold of *p*<5×10^−8^ (--gwas-thresh 5e-8) and an r^2^ threshold of 0.1 (--clump-r2 0.1). In discovery analyses to identify potentially causal metabolites for BPD, we set the minimum number of significant variants required for GSMR analysis to 10 (--gsmr-snp-min 10). In follow-up and replication analyses, we set this parameter to 1 (--gsmr-snp-min 1). To protect against horizontal pleiotropy, we specified a *p*-value threshold of 0.01 for HEIDI-outlier variant filtering (--heidi-thresh 0.01) (7). To estimate variant correlations, we utilized an individual-level reference genotype panel comprising 4,994 participants from the INTERVAL population study (16). For the discovery analyses, we set an adjusted significance threshold of *p*_adj_ <0.05/913 = *p*_adj_ <5.48×10^−5^ by applying a Bonferroni correction for the total number of metabolites tested. We plotted the GSMR estimates using the forestplot (v3.1.1) (https://CRAN.R-project.org/package=forestplot) and corrplot (v0.92) (17) R packages. We performed a ‘leave-*FADS*-locus-out’ analysis by first removing variants at the *FADS* cluster (chr11:60567097-62659006, hg19) from the exposure input file, and then re-running GSMR using the same parameters as above with the minimum number of significant variants set to 1 (--gsmr-snp-min 1). We defined the start position for the *FADS* cluster as the 5’ *FADS1* hg19 coordinate minus 1Mb (i.e., 61567097-1Mb) and the end position as the 3’ *FADS3* hg19 coordinate plus 1Mb (i.e., 61659006+1Mb).

### Sensitivity analyses

To check the sensitivity of our significant (*p*_adj_ <5.48×10^−5^) GSMR findings to alternative MR methods, we performed inverse-variance weighted Mendelian randomization (IVW-MR) and Mendelian randomization-Egger (MR-Egger) using the MendelianRandomization R package (v0.70) (18). As input, we used the exposure and outcome summary data for the same genetic instruments selected by GSMR for each of the 33 metabolites as appropriate. Since we used an r^2^ threshold of 0.1 for the GSMR LD clumping procedure, we accounted for any residual LD between instruments by supplying a variant correlation matrix generated in plink (v1.07) using the above-mentioned INTERVAL reference genotype panel.

### Replication analyses

We performed replication analyses using summary data from an earlier multi-trait GWAS of 529 metabolites quantified in an independent sample of 7,824 European individuals (14). Since the metabolites from both the Surendran et al. (2022) (13) and Shin et al. (2014)(14) studies were annotated with Metabolon IDs, we used these IDs and the metabolite names to identify unambiguously matching metabolites from the set of 33 significant metabolites highlighted in the discovery GSMR analysis. Overall, three of the 33 metabolites matched unambiguously, one of which we excluded due to a lack of any genome-wide significant instruments (1-arachidonoyl-GPE [20:4n6]). We performed GSMR analyses using the remaining two metabolites from Shin et al. (14) as exposures and BPD from Mullins et al. (2021) (5) as the outcome variable.

### Distance-based clumping of instruments

We first extracted all the 293 unique genetic variants used to instrument at least one of the 33 significant metabolites in our main GSMR analyses. We ordered them by chromosome and position, used the first variant to initiate the first clump, and then sequentially checked the distance between the next variant and the preceding one. If the distance was <500Kb, we added the variant to the current clump; and if the distance was ≥500Kb, we used that variant to initiate the next clump. We continued this process until all variants were assigned to a clump. We annotated all instruments using the Variant Effect Predictor (VEP) (GRCh37, release 109) (19) restricting results to show only a single consequence per variant, selected according to the criteria outlined at: https://asia.ensembl.org/info/docs/tools/vep/script/vep_other.html#pick.

### Statistical colocalization

We conducted systematic pairwise statistical colocalization using the coloc.abf function from the Coloc R package (v5.1.0.1) (20), with priors and parameters set to default. The Coloc method has been described in detail previously, but briefly, it is a suite of Bayesian tools that takes locus-specific summary data for a pair of traits as input. The coloc.abf function returns posterior probabilities (PPs) to estimate the likelihood that five distinct scenarios – or hypotheses – are true, assuming a single causal variant for each trait. These five hypotheses are: H_0_, there is no causal variant for either trait at the locus; H_1_/H_2_, there is a causal variant for the first/second trait only; H_3_, there are two distinct causal variants at the locus, one for each trait; H_4_, both traits share the same causal variant. We extracted all available summary statistics at the *FADS* cluster (chr11:60567097-62659006, hg19) for BPD and each of the 33 metabolites. We used the same start and end coordinates to define the *FADS* cluster as for the leave-locus-out analysis.

### Phenome-wide association study (PheWAS)

We performed a PheWAS of rs174592 using the PheWAS webtool available at the Integrative Epidemiology Unit (IEU) OpenGWAS project. We downloaded a .csv file containing the results and filtered the output as follows: (i) we discarded traits/datasets with *p*>5×10^−8^; (ii) we removed all traits/datasets derived from high-throughput molecular trait panels (e.g., trait IDs starting with “eqtl-”, “pqtl-”, and “met-”); and (iii) for traits represented by more than one dataset, we kept the dataset with the lowest *p*-value.

### Mendelian randomization of lipid fractions and lipoprotein exposures

We used the tophits() function from the ieugwasr R package (v0.1.5) to select independent genetic instruments for the lipid fraction and lipoprotein traits. The tophits() function incorporates a linkage disequilibrium clumping procedure; we used the default clumping parameters (i.e., r2<0.001, distance>10Mb). We then extracted the matching BPD GWAS summary data, performed harmonization to ensure the estimates from both the lipid and BPD data were oriented to the same allele, and then ran IVW-MR using the MendelianRandomization R package (v0.70) (18) with the lipid traits as exposures and BPD as outcome.

## Results

### Dysregulation of the arachidonic acid synthesizing pathway is a potential risk factor for bipolar disorder

To identify potential causal plasma metabolites for BPD, we conducted exploratory MWMR analyses. After Bonferroni correction for 913 tests (*p*<5.48×10^−5^), these analyses highlighted 33 metabolites with evidence of a link with BPD risk (**Figure 1A**). Sensitivity analyses (see **Methods and Materials**) yielded consistent estimates (**Figure S1, Table S2**), and we did not observe any significant evidence of reverse causality after Bonferroni correction for 33 tests (*p*<0.002) (**Table S3**). To validate our results, we conducted replication analyses using summary data from an earlier metabolomic study(14). Due to differences in metabolomic coverage between the discovery and replication datasets, we could only perform replication analyses for 2 of the 33 metabolites (see **Methods and Materials**). Nevertheless, for both metabolites we observed highly significant estimates in the same direction as in the discovery analyses (**Figure 1B**).

**Figure 1:**
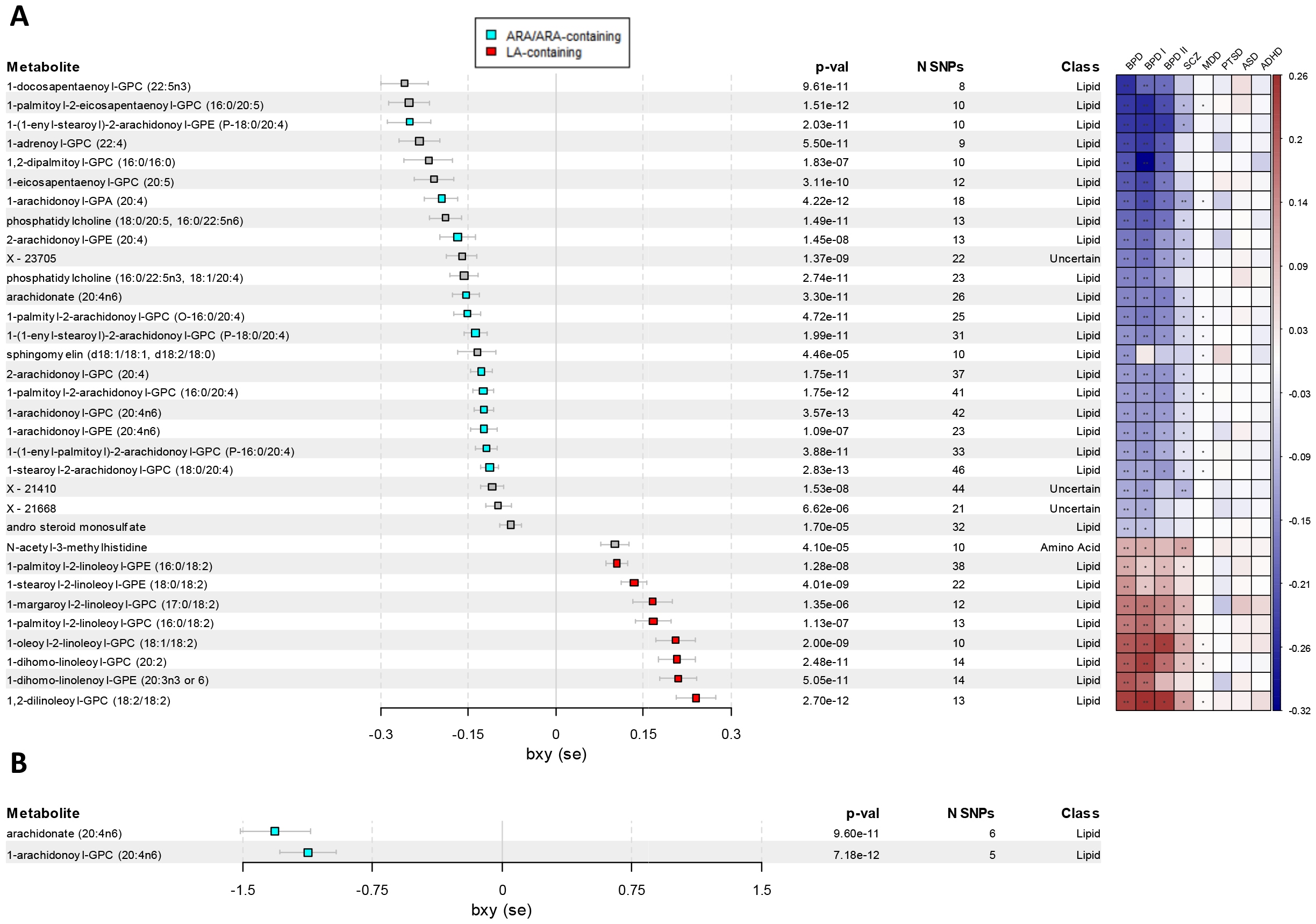
Metabolome-wide Mendelian randomization reveals 33 metabolites with significant (*p*<5.48×10^−5^) associations with bipolar disorder. **(A)** Plots depicting the bxy (which approximates the logOR) estimates of the slope (effect size) of the relationship between each of the 33 significant metabolites from Surendran et al. (2022) and bipolar disorder (see forest plot, left), as well as bipolar disorder along with the two main bipolar disorder subtypes and several other psychiatric disorders (see heatmap, right). **(B)** Plot depicting the bxy (logOR) estimates of the causal effect on bipolar disorder of two of the 33 metabolites that we could unambiguously match from an earlier metabolomic study by Shin et al. (2014) used here for replication analyses. The number of approximately independent instruments (r2<0.1) are indicated in the N SNPs column. ARA: arachidonic acid, LA: linoleic acid, BPD: bipolar disorder, BPDI: BPD type 1, BPDII: BPD type 2, SCZ: schizophrenia, MDD: major depressive disorder, PTSD: post-traumatic stress disorder, ASD: autism spectrum disorder, ADHD: attention deficit hyperactivity disorder. ** *p*_*adj*_<5.48×10^−5^, * *p*<0.05

The vast majority of the 33 significant metabolites were lipids (29/33, 88%), one was an amino acid, and the remaining three were either unannotated or uncertain. Notably, we found that arachidonic acid, a polyunsaturated omega-6 fatty acid, was associated with BPD risk in both its free form and as a side chain of 11 different complex lipids (**Figure 1A**). Importantly, all estimates for ARA and ARA-containing lipids consistently showed that lower levels of these metabolites were associated with higher BPD risk. Conversely, we also found eight significant linoleic acid (LA)-containing lipids, all with estimates in the opposite direction (**Figure 1A**). Since ARA is synthesized through the desaturation and elongation of dietary linoleic acid (LA) (21), this pattern of opposing estimates suggests a potential causal link between ARA synthesizing mechanisms and BPD.

### The identified metabolite associations are specific to BPD

To assess the specificity of the observed metabolic associations, we performed GSMR to derive estimates for several related psychiatric outcomes (see **Methods and Materials**). We found broadly similar estimates for BPD type I and II (**Figure 1A**). Although larger *p*-values were generally observed for BPD type II, this was likely due to the smaller number of cases available for type II (up to 6,781) relative to type I (up to 25,060). This suggests that the observed causal associations were not specific to either BPD subtype. We also found several metabolite associations with schizophrenia and depression, although these associations were only nominally significant in general (*p*<0.05) and exhibited much smaller effect sizes compared to BPD (**Figure 1A**). These findings support the notion of a BPD-specific role for these metabolites.

### BPD-associated metabolites may mediate a favourable response to lithium treatment

To determine whether the 33 metabolites might also mediate the response to lithium treatment, we conducted GSMR analyses utilizing summary statistics from a GWAS of lithium response conducted by ConLi^+^Gen. (2) Overall, we found nominal significance (*p*<0.05) for the estimates of only two metabolites for lithium response represented as a continuous variable (see **Methods and Materials**) (**Figure 2A**). However, when we directly compared the estimates for BPD and lithium response, we observed a highly significant inverse correlation with the continuous phenotype (*r*=−0.78, *p*=1.41×10^−7^) and a nominally significant inverse correlation with the dichotomous (i.e., responders vs. non-responders) phenotype (*r*=−0.48, *p*=0.005) (**Figure 2B**). These findings suggest that metabolites associated with lower BPD risk, like ARA, may also be associated with a higher likelihood of a favourable response to lithium.

**Figure 2:**
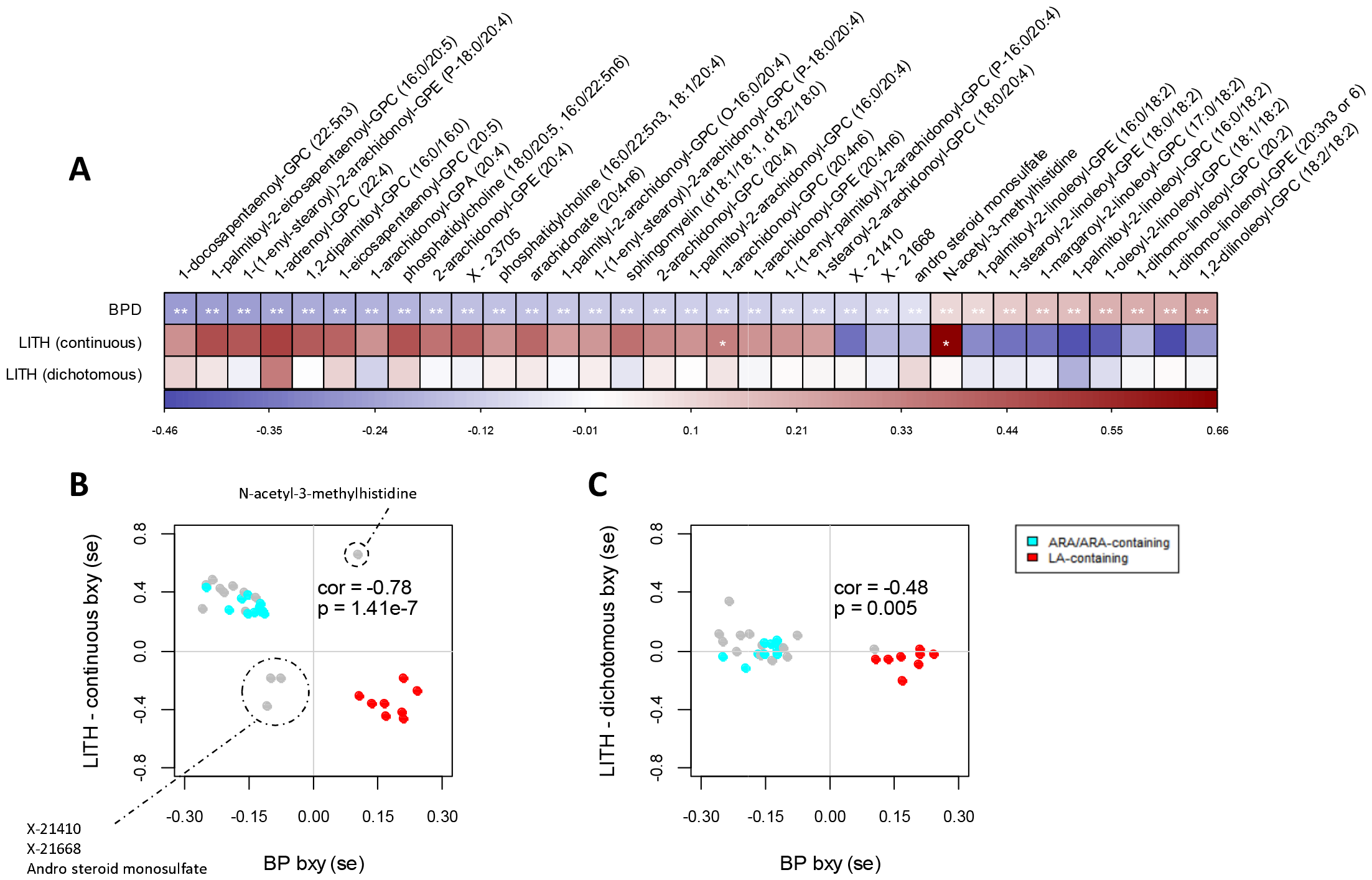
Metabolites associated with lower risk of bipolar disorder may also be associated with a higher likelihood of a favourable response to lithium treatment. **(A)** Heatmap plotting the bxy (logOR) estimates from GSMR analyses for each metabolite for bipolar disorder and lithium response. ** *p*_*adj*_ <5.48×10^−5^, * *p*<0.05. **(B, C)** Scatterplots comparing the bipolar disorder and **(B)** continuous and **(C)** dichotomous lithium response bxy (logOR) estimates from GSMR analyses. The Pearson’s *r* correlation and associated *p*-value are indicated in the plot. ARA: arachidonic acid, LA: linoleic acid, BPD: bipolar disorder, LITH: lithium response.

### Lipid associations with BPD are driven primarily by the FADS1/2/3 locus

Next, we explored the mechanisms driving the observed associations with BPD. We found that almost 40% (110 of 293) of the GSMR-selected instruments reside at the *FADS1/2/3* cluster (chr11q12.2) (**Table S4**), which encodes a family of fatty acid desaturase (*FADS*) enzymes responsible for converting LA into ARA.(21) We performed a ‘leave-*FADS*-locus-out’ analysis by removing all 110 instruments at the locus (chr11:60567097-62659006, hg19; see **Methods and Materials**) and re-running GSMR with the 33 metabolite exposures. As a result, only 4 of the 33 metabolites retained a signal for BPD at the previously defined significance threshold (*p*<5.48×10^−5^) (**Table S5**), indicating that the observed metabolite associations were driven primarily by the *FADS1/2/3* cluster.

Accordingly, a robust genome-wide significant signal for BPD resides at the *FADS1/2/3* cluster.(5) The minor G allele of the sentinel variant, rs174592, is associated with a higher risk of BPD and lower levels of plasma ARA, which is consistent with the directionality observed in our GSMR analyses (**Figure 1A**). To determine whether the 33 associated metabolites share the same genetic aetiology with BPD at the *FADS1/2/3* cluster, we performed statistical colocalization analyses for each BPD-metabolite pair. Overall, we found robust evidence of colocalization (i.e., PP_H4_>0.8, see **Methods and Materials**) for 27 of the 33 metabolites (**Figure 3**), indicating that most of the metabolites do indeed share the same genetic aetiology with BPD at the *FADS1/2/3* cluster. This includes ARA and all the ARA-containing complex lipids, as well as all but one of the LA-containing lipids.

**Figure 3:**
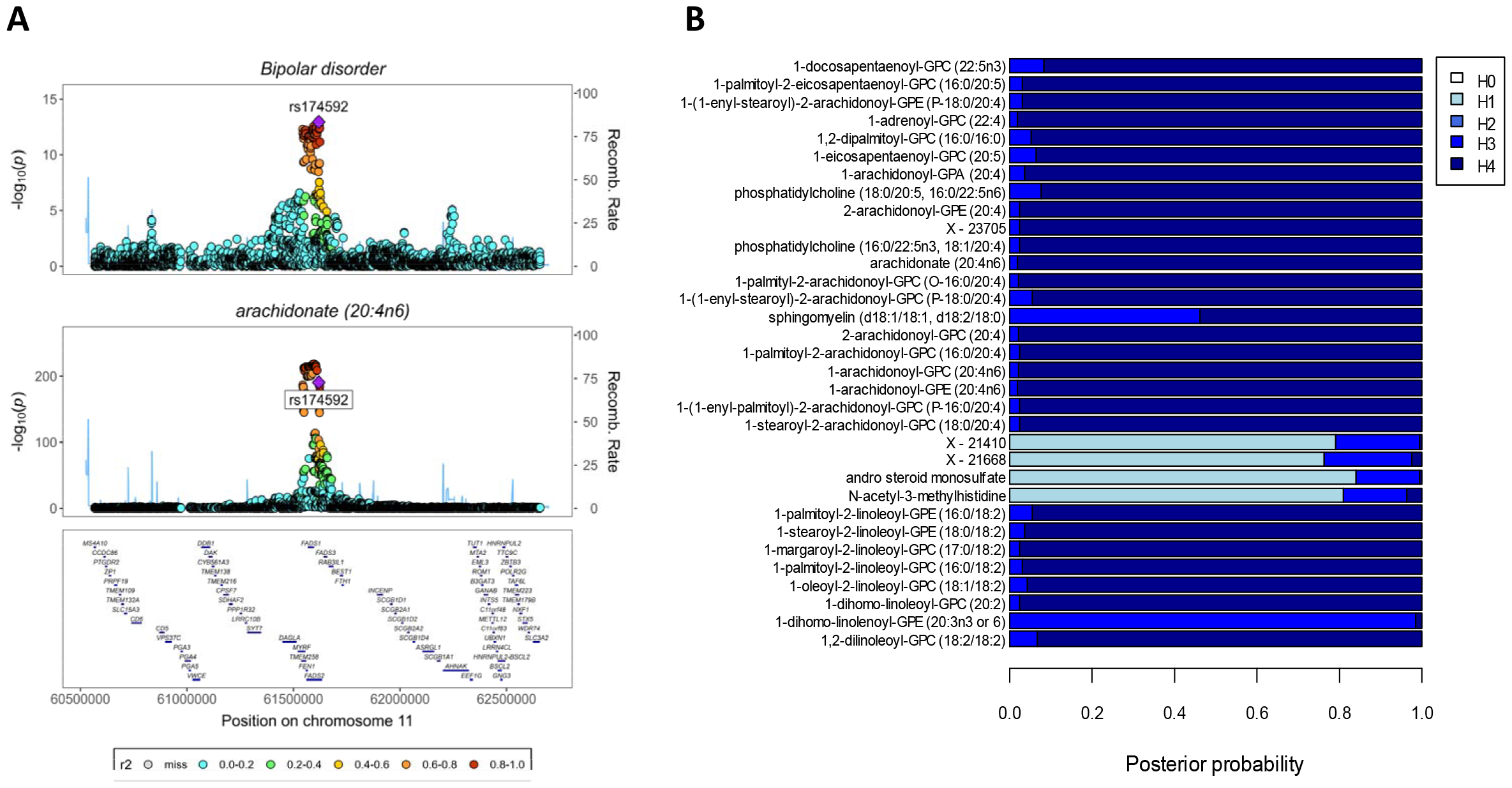
The genome-wide significant bipolar disorder signal at the *FADS1/2/3* cluster shares the same genetic aetiology with 27 of the 33 metabolites tested, including arachidonic acid. **(A)** Regional association plot centred on the *FADS1/2/3* locus depicting the bipolar disorder (top) and arachidonic acid (bottom) signals. The bipolar disorder sentinel variant, rs174592, is indicated. **(B)** Stacked bar plot depicting posterior probabilities (PP) of H0 (no causal variant), H1 (causal variant for bipolar disorder only), H2 (causal variant for metabolite only), H3 (two distinct causal variants), and H4 (one shared causal variant) returned by Coloc. We consider PP_H4_>0.8 as evidence of shared genetic aetiology.

### No evidence of a causal effect of major lipid fractions or lipoproteins on BPD risk

The *FADS1/2/3* cluster is a pleiotropic locus and is known to play a critical role in the regulation of major lipid fractions such as low-density lipoprotein (LDL) and triglycerides. Thus, to assess whether the *FADS1/2/3* cluster might instead be exerting its effect on BPD risk through the regulation of broad lipid fractions or lipoproteins, we first performed a phenome-wide association study (pheWAS) of the BPD sentinel variant using the openGWAS resource to select potentially causal lipid traits. We then followed this up by performing two-sample MR analyses.

The overall pheWAS output after filtering (see **Methods and Materials**) is presented in **Table S6**. We extracted all major lipid fraction and lipoprotein datasets associated with rs174592 at genome-wide significance (*p*<5×10^−8^). This left us with 7 unique lipid traits/datasets (**Table S6**): high density lipoprotein (HDL) cholesterol, triglycerides, apolipoprotein B, LDL cholesterol, total cholesterol, apolipoprotein A-I, and apolipoprotein A. We selected independent (*r*^2^<0.001) instruments for these 7 lipid datasets by LD clumping, and then performed a series of inverse-variance weighted MR analyses with BPD as the outcome variable. We did not find evidence (i.e., *p*>0.05) of a potential causal role for any of these broad lipid traits on BPD (**Table S7**), which is consistent with the *FADS1/2/3* cluster exerting its effect on BPD risk through more specific and more proximal downstream mediators, such as ARA synthesis.

## Discussion

This two-sample MWMR study revealed 33 circulating metabolites associated with BPD, most of them lipids. Chief among these metabolites was ARA in its free form along with several complex lipids containing either an ARA or LA sidechain. We showed that most of these metabolite associations were driven by genetic instruments at the *FADS1/2/3* cluster, which encodes a family of fatty acid desaturase enzymes responsible for the desaturation of omega 6 PUFAs in the LA to ARA pathway as well as omega 3 PUFAs in the αLA to eicosapentaenoic acid (EPA) to docosahexaenoic acid (DHA) pathway (22). Taken together, these findings suggest that the conversion of LA into ARA by the *FADS1/2/3* cluster genes may play a key role in BPD aetiology.

A potential role for PUFAs in BPD has been widely posited over the past decade, though attention has primarily been focused on omega-3 PUFAs such as EPA and DHA (22). A key finding from this two-sample MWMR study suggests that increased synthesis of ARA, an omega-6 PUFA, may lower BPD risk. Although the ARA pathway has previously been implicated in the pathophysiology of BPD (23), to our knowledge, ours is the first study to highlight a potential causal role. Further work is required to uncover the relevant downstream mechanisms, though studies have shown that ARA plays a vital role in the central nervous system (CNS), both as a major phospholipid constituent of neuronal and glial cell membranes and as a signalling molecule (24).

Given its presence in human milk, ARA is considered essential for infant brain development and is added to infant formulas in many countries (25). ARA may therefore exert an effect on BPD risk by affecting neurodevelopmental pathways, which would be consistent with contemporary views of BPD as a neurodevelopmental disorder (26). In children and adults, ARA can be sourced either directly from meat and seafood products, or indirectly by de novo synthesis from dietary LA (e.g., nuts, seeds, oils) via the *FADS1/2/3* cluster genes (21). Conversely, infants lack the ability to synthesize ARA from LA, and so are completely reliant on human milk or formula for their ARA intake (27). Future studies to assess ARA supplementation as a potential preventative strategy for BPD may therefore be warranted, particularly in children or infants with poor natural dietary sources of ARA.

A GWAS of fatty acid levels in breast milk from >1,000 Bangladeshi mothers revealed that the *FADS1/2/3* cluster was the primary driver of fatty acid levels, and that of the 33 fatty acids measured, ARA was the primary fatty acid influenced by this gene cluster (28). Specifically, the authors found that the sentinel variant at the locus, rs174556, was associated with a per major allele effect size of 17% higher ARA levels (28). This is a large effect, and so lower levels of ARA in human milk due to genetic variation at the *FADS1/2/3* cluster could play a role in modulating BPD risk. Indeed, the sentinel variant associated with ARA in breast milk is in moderate to high linkage disequilibrium with the BPD sentinel, rs174592 (r^2^=0.67, 1000G EUR), suggesting the two signals do overlap. However, if the levels of ARA in breast milk were a driver of the BPD signal, then the mothers’ genotype at the FADS1/2/3 cluster would be more important than the infants’. Thus, a hypothetical cross-generational effect like this would suggest the *FADS1/2/3* cluster exerts a larger effect on BPD risk than current data suggest.

In this study we have shown that the BPD GWAS signal at the *FADS1/2/3* cluster is likely driven by PUFA metabolism, suggesting ARA may be the primary effector. Although the liver is generally considered to be the central organ of PUFA metabolism, the *FADS1/2/3* cluster genes are expressed widely across human tissues and cell types, including in the brain (29). It is therefore unclear whether this cluster impacts BPD risk via distal mechanisms either in the liver or another peripheral tissue, or by local activity in the brain. A recent study using a *Fads1/2* knockout mouse model of BPD showed that peripheral, but not central, *Fads1/2* gene knockout was necessary to recapitulate the BPD-like phenotype, as evidenced by a loss of the phenotype after conditional knockout in the brain (30). Indeed, ARA and other PUFAs are actively transported into the brain across the blood-brain-barrier (31), and recent GWASs have uncovered genome-wide significant associations between variants at the *FADS1/2/3* cluster and several brain imaging derived phenotypes including cortical thickness and surface area (32, 33). Taken together, this is all consistent with peripheral PUFA metabolism having the potential to impact central mechanisms.

A major clinical implication of the associations observed here between circulating metabolites and BPD relates to the search for biomarkers. There are currently no clinically approved psychiatric biomarkers, though predictive modelling has been widely applied in attempts to identify biosignatures of different psychiatric disorders. For example, a recent study identified a lipid signature for schizophrenia that also translates to both BPD and major depressive disorder (34). High resolution profiling of circulating PUFAs in psychiatric patients may therefore prove to be crucial in facilitating psychiatric biomarker discovery.

There are several potential limitations to this study. First, although we observed an inverse correlation between the BPD and lithium response GSMR estimates across the 33 BPD-associated metabolites, the lithium response estimates were based on a GWAS of just 2,039 BPD patients (2). A potential role for these metabolites in modulating lithium response should therefore be considered suggestive at best, though several previous studies have shown that lithium and other mood stabilizers impact the ARA pathway in animals (35, 36). Second, it is unclear to what extent patients from the BPD GWAS by Mullins et al. utilised in this study had been prescribed lithium. Although MR studies are more robust to confounding from medication and other lifestyle factors than observational studies, it is possible that the signals detected in our study reflects a lithium-responsive BPD subtype. Further studies to explore this in more deeply phenotyped participants are therefore needed.

Third, since our findings were based on GWASs of European participants, the extent to which they are applicable to other populations is unclear. However, the GWAS signal for BPD at the *FADS1/2/3* cluster was originally identified in a Japanese population (37), suggesting the relevance of our findings likely does extend beyond Europeans. Fourth, although we found little to no evidence to support a role for these 33 BPD-associated metabolites in other related psychiatric disorders, it is possible that they may be relevant for certain psychiatric subtypes. For example, the depression phenotype used in the Howard et al. GWAS was very broad, and so MR studies utilising future GWASs focused on specific depression subtypes that adhere to more uniform and clinically relevant diagnostic criteria may yield different results.

Finally, our replication efforts were hampered by differential metabolomic coverage between the metabolomic discovery and replication datasets. Future studies should therefore focus primarily on replication and validation. Indeed, studies applying the Metabolon HD platform and other high-resolution platforms to independent population cohorts will be crucial to enable comprehensive replication analyses. Moreover, the application of platforms with ever-increasing coverage of PUFAs will help to pinpoint the most relevant metabolites involved in BPD aetiology. We propose that future validation studies should utilise the *Fads1/2* knockout mouse model of BPD (30) discussed earlier. These mice may represent a suitable preclinical model to assess the efficacy of PUFA supplementation (including ARA) in treating BPD-related symptoms, thereby highlighting specific PUFAs for subsequent randomised clinical trials.

In conclusion, our study suggests that higher levels of ARA may reduce the risk of BPD. Preclinical models and randomized clinical trials are needed to rigorously assess a potential role for ARA supplementation (and other PUFAs) in facilitating BPD prevention and treatment, particularly in individuals carrying BPD risk alleles at the *FADS1/2/3* cluster. More broadly, our findings also support potential avenues for precision health interventions focused on early life nutrition to ensure that infants and children are receiving enough ARA and other PUFAs to support optimal brain development, which may also reduce the risk of BPD.

## Data Availability

All data produced in the present study are available upon reasonable request to the authors

https://omicscience.org/apps/mgwas/

https://pgc.unc.edu/for-researchers/download-results/

https://ftp.ebi.ac.uk/pub/databases/gwas/summary_statistics/GCST012001-GCST013000/GCST012487/

## Acknowledgements

We thank Dr. Praveen Surendran for providing access to the complete summary data for the 913 metabolite exposures we tested. We also thank the Psychiatric Genetics Consortium, the IEU OpenGWAS project, and the NHGRI-EBI GWAS catalog for providing access to the complete summary data for the outcomes we tested.

## Conflicts of Interest

None to declare.

## Notes

### Competing Interest Statement

The authors have declared no competing interest.

### Funding Statement

This study did not receive any funding

### Author Declarations

All source data were publicly available prior to the initiation of the project. The data are available at: (1) https://omicscience.org/apps/mgwas/, (2) https://pgc.unc.edu/for-researchers/download-results/, (3) https://ftp.ebi.ac.uk/pub/databases/gwas/summary_statistics/GCST012001-GCST013000/GCST012487/

