## Supplementary material for "A metabolome-wide Mendelian randomization study identifies dysregulated arachidonic acid synthesis as a potential causal risk factor for bipolar disorder": Figure S1

### Slide 1
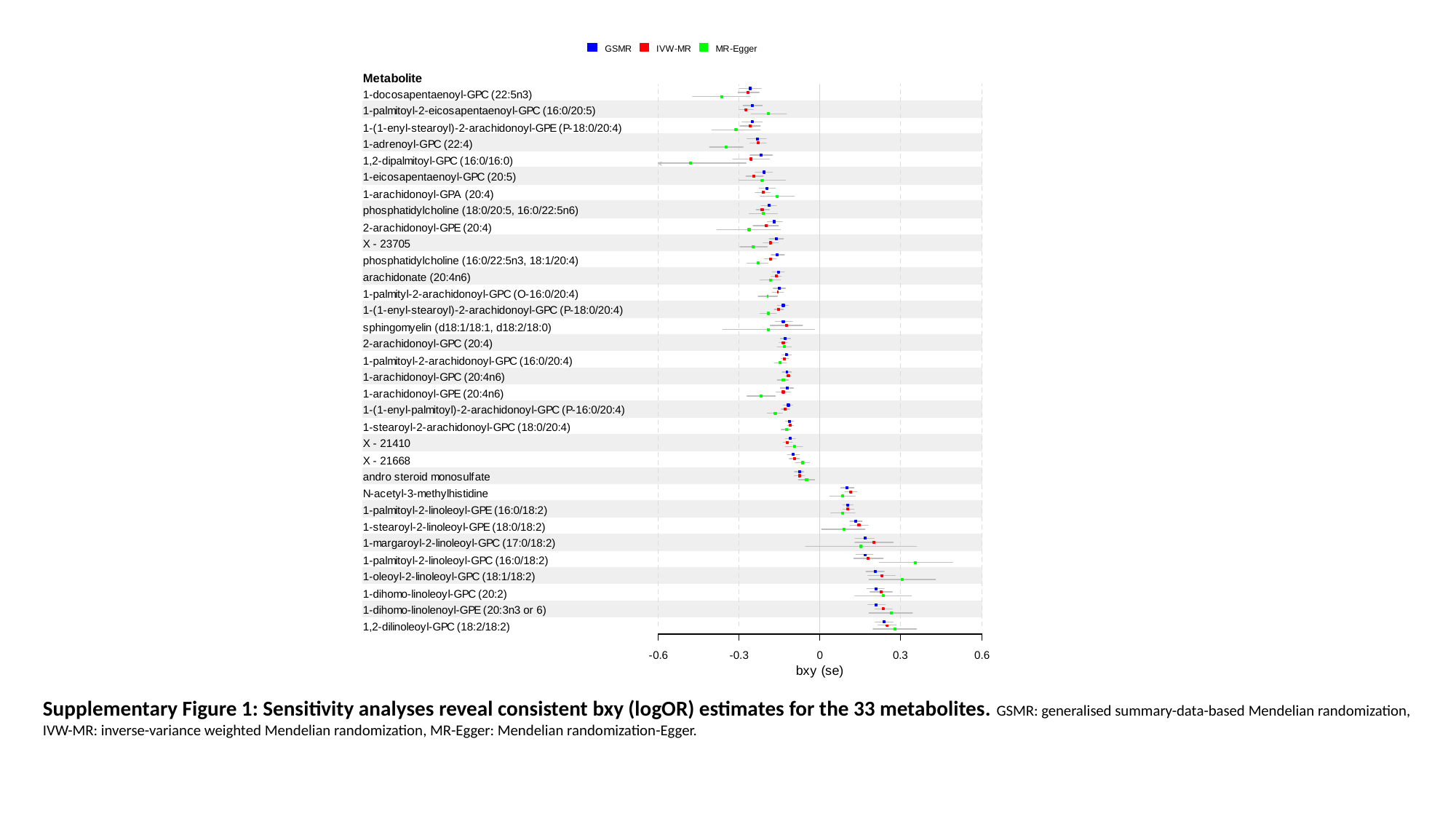

Supplementary Figure 1: Sensitivity analyses reveal consistent bxy (logOR) estimates for the 33 metabolites. GSMR: generalised summary-data-based Mendelian randomization, IVW-MR: inverse-variance weighted Mendelian randomization, MR-Egger: Mendelian randomization-Egger.
