## Supplementary Methods for "A metabolome-wide Mendelian randomization study identifies dysregulated arachidonic acid synthesis as a potential causal risk factor for bipolar disorder"

*GWAS summary statistics*. For this study we utilized summary data from:

1. A multi-trait GWAS of 913 plasma metabolites (**Table S1**) quantified in 14,296 European individuals using the Metabolon HD4 platform (1). For replication, we utilized an earlier multi-trait GWAS of 529 plasma or serum metabolites quantified in 7,824 European individuals using liquid-phase chromatography and gas chromatography separation coupled with tandem mass spectrometry (2). Overall, there were 219 metabolites with matching Metabolon IDs shared between the discovery and replication datasets. Moreover, the discovery and replication data were generated in independent cohorts of European ancestry. Briefly, the discovery data were from two UK-based cohorts, INTERVAL (mean age 44 years, 48.8% female) and EPIC-Norfolk (mean age 59.8 years, 53.3% female), while the replication data were from TwinsUK (mean age 53 years, 93% female) and KORA (mean age 61 years, 50% female), a UK- and a Germany-based cohort, respectively.
2. The most recent and largest (at the time of writing) genome-wide association studies available from the Psychiatric Genetics Consortium for: (i) bipolar disorder (BPD) in up to 413,466 Europeans, including 41,917 cases (3); (ii) schizophrenia (SCZ) in up to 130,644 Europeans, including 53,386 cases (4); (iii) broadly-defined depression in up to 500,199 Europeans, including 170,756 cases (5); (iv) attention deficit hyperactivity disorder (ADHD) in up to 186,843 Europeans, including 38,691 cases (6); (v) anorexia nervosa (AN) in up to 55,525 Europeans, including 16,992 cases (7); (vi) autism spectrum disorder (ASD) in up to 46,350 Europeans, including 18,381 cases (8); and (vii) post-traumatic stress disorder (PTSD) in up to 151,309 Europeans, including 23,185 cases (9).
3. A GWAS of response to lithium treatment in BPD patients conducted by the international consortium on Li^+^ genetics (ConLi^+^Gen). Lithium response was retrospectively quantified using the ALDA scale (10) and GWASs were conducted with lithium response represented on a continuous scale (i.e., score between 0-10, with higher scores indicating a better response) and as a dichotomous variable (i.e., score ≥7: responder; score <7: non-responder). We utilised summary data generated using both the continuous (2,039 European BPD cases) and dichotomous (2,343 European BPD cases) variables (10). Both datasets were downloaded from the NHGRI GWAS catalog.
4. GWASs of major lipid fractions and lipoproteins – all measured in European individuals – accessed through the Integrative Epidemiology Unit (IEU) openGWAS project. The traits and datasets (represented here by openGWAS codes) accessed were as follows: high density lipoprotein (HDL) cholesterol (ieu-b-109) (11); triglycerides (ieu-b-111) (11); apolipoprotein B (ieu-b-108) (11), low density lipoprotein (LDL) cholesterol (ebi-a-GCST90002412) (12); total cholesterol (ukb-d-30690_raw) (<http://www.nealelab.is/uk-biobank/>); apolipoprotein A-I (ieu-b-107) (11); and apoliporotein A (ukb-d-30630_raw) (<http://www.nealelab.is/uk-biobank/>).

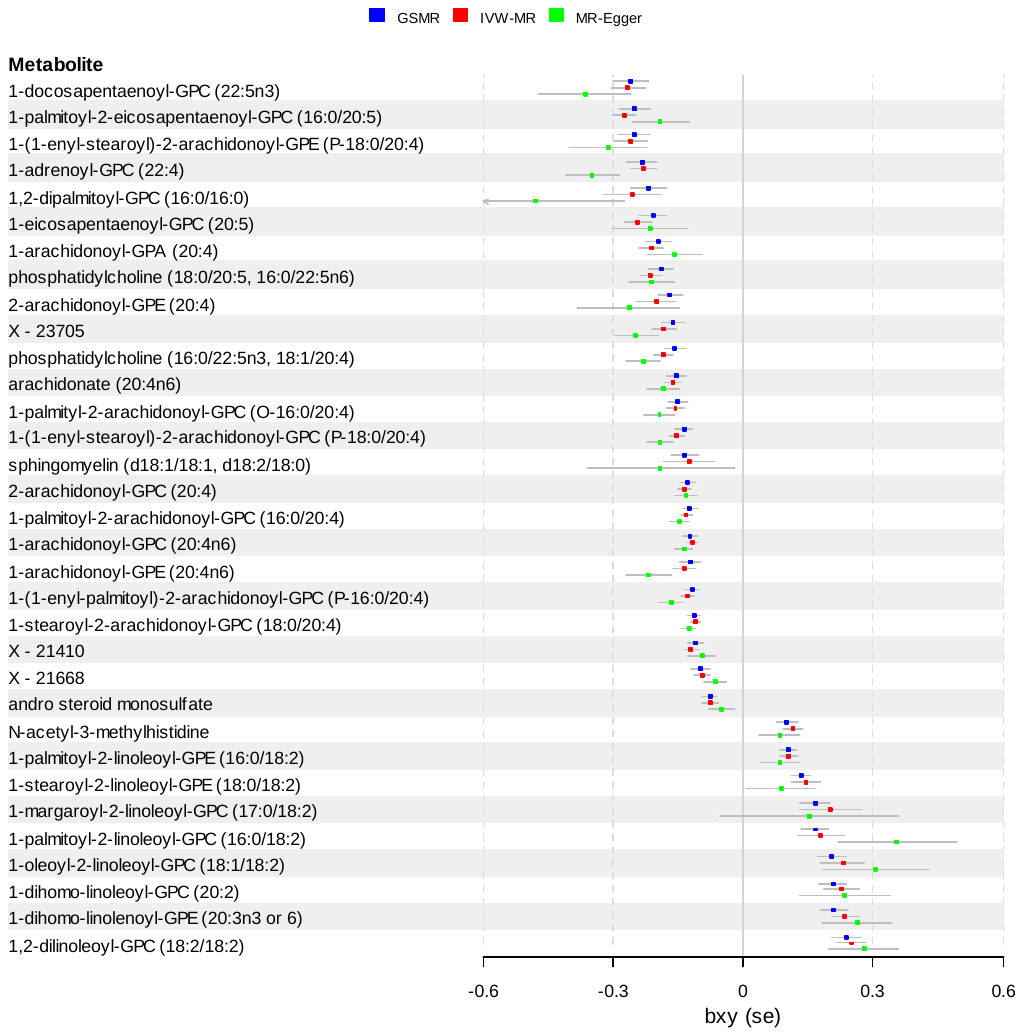

**Supplementary Figure 1: Sensitivity analyses reveal consistent bxy (logOR) estimates for the 33 metabolites.** GSMR: generalised summary-data-based Mendelian randomization, IVW-MR: inverse-variance weighted Mendelian randomization, MR-Egger: Mendelian randomization-Egger.
